# Acupuncture for emotional disorder and quality of life in patients with polycystic ovary syndrome-A protocol for a systematic review and meta-analysis

**DOI:** 10.1101/2022.10.10.22280920

**Authors:** Chunping Zhang, Yiwen Zhang, Jian Chen, Chuanzhu Yan

**Author notes:** Correspondence:Chunping Zhang, <u>No.16369 qianfoshan Campus, Shandong University of Traditional Chinese Medicine</u>. Contributions:(I) Conception and design: C Zhang, Y Zhang, J Chen, C Yan; (II) Administrative support: None; (III) Provision of study materials or patients: None; (IV) Collection and assembly of data: C Zhang, Y Zhang, C Yan; (V) Data analysis and interpretation: C Zhang, Y Zhang, J Chen; (VI) Manuscript writing: All authors; (VII) Final approval of manuscript: All authors. All authors contributed to this work.

## Abstract

**Background:** Infertility, obesity, hairiness, acne and other symptoms of polycystic ovary syndrome can easily lead to emotional disorders, such as anxiety and depression, and also affect the patients’ quality of life. A large number of studies have shown that acupuncture has a good effect on polycystic ovary syndrome, but there is no evidence that acupuncture can relieve the affective disorders and improve the quality of life in patients with polycystic ovary syndrome.

**Objectives:** To assess the effectiveness and safety of acupuncture for emotional disorder and quality of life in patients with PCOS.

**Methods and analysis:** From inception to July 31, 2022, a total of four English-language databases (PubMed, Embase, Web of science and the Cochrane Central Register of Controlled Trials) and four Chinese-language databases (the China National Knowledge Infrastructure, Wanfang, VIP, Chinese Biomedical) were searched. The randomized clinical trial (RCT) of acupuncture for polycystic ovary syndrome was selected for inclusion in the study. Statistical analysis will be performed by using the Cochrane Review Manager (RevMan 5.4) software. The strength of the evidence will be assessed according to the Grading of Recommendations Assessment, Development, and Evaluation (GRADE).

**Results and conclusion:** This study will systematically evaluate the efficacy and safety of acupuncture for emotional disorder and quality of life in patients with polycystic ovary syndrome.

## Introduction

Polycystic ovary syndrome (PCOS) is one of the most common endocrine diseases in women of reproductive age, characterized by abnormal ovulation function and excessive androgen secretion[1,2],which clinical manifestations are menstrual disorder, hirsutism, acne, obesity, infertility, etc. The incidence rate of PCOS is 6%∼ 10%, even as high as 15%[3,4]. The 2018 international evidence-based guidelines pointed out that the psychological problems of PCOS patients include anxiety, depression, sexual fear, sexual apathy and other sexual psychological disorders, as well as eating disorders[5],the most common of which is anxiety and depression. Studies have shown that PCOS patients have anxiety and depression of different severity[6]. According to statistics, about 60% of PCOS patients have emotional problems such as anxiety and depression, and the incidence rate of anxiety and depression is 3-5 times than that of healthy people[7-9]. The causes of anxiety and depression in PCOS patients are not very clear. It is generally believed that they may be related to PCOS symptoms (obesity, hairiness, acne, infertility, etc.), excessive androgen secretion or other factors [10-12]. However, some studies have shown that high BMI or hyperandrogenism is not related to the symptoms of anxiety or depression[8]. Negative emotions can aggravate the symptoms of PCOS patients. Studies have shown that anxiety and depression can reduce the PCOS patients’quality of life, which is the largest factor leading to low quality of life[13,14]. In addition, anxiety and depression can also damage the brain function of PCOS patients and increase the risk of eating disorders and sexual dysfunction[15-16]. Therefore, more attention and intervention should be given to the emotional state of PCOS patients. The guidelines recommend that the first-line treatment for PCOS is oral contraceptives, which are not effective in alleviating anxiety and depression.

Complementary and alternative medicine(CAM) has become a choice of many PCOS patients[17]. Acupuncture is one of CAM, which has advantages of green, safe, convenient and no side effects. There is evidence that acupuncture can alleviate the symptoms of PCOS as well as the anxiety and depression associated with it[18]. However, some research show that acupuncture may aggravate the PCOS patients’anxiety [19,20]

At present, most of the domestic and foreign literature reports on acupuncture are mainly aimed at the physical symptoms or changes in biochemical indicators of PCOS patients, while psychological symptoms are often ignored. Although there have been many meta-analysis articles on acupuncture improving PCOS related symptoms[21,22], there is no systematic review on acupuncture improving emotional disorders of PCOS. In this study, we will summarize the evidence from randomized controlled trials (RCTs) to evaluate the effectiveness and safety of acupuncture in alleviating emotional symptoms in PCOS patients.

## 2. Method

### 2.1 Study registration

The systematic review protocol has been registered in the International Prospective Register of Systematic Reviews (PROSPERO) (Registration number:CRD42022357372). This protocol is reported in line with the Preferred Reporting Items for Systematic Review and Meta-Analysis Protocols (PRISMA-P) statement guidelines [23].

### 2.2 Study criteria

#### 2.2.1 Types of studies: Randomized controlled trials (RCTs) of acupuncture therapy

for emotional disorder and quality of life in patients with PCOS will be included. Summary results of ongoing and completed trials published on the clinical trial registration platform will also be included. This systematic review will exclude non-randomized controlled trials, retrospective studies, case reports, non-controlled trials, and animal studies.

#### 2.2.2 Types of participants

Participants who were diagnosed with PCOS according to the European Society of Human Reproduction and Embryology (ESHRE) and the American Society of Reproductive Medicine (ASRM)consensus in Rotterdam in 2003, will be included regardless of their age, race, and background. [24]

#### 2.2.3 Types of interventions

##### Experimental intervention

Acupuncture is the stimulation of acupuncture points by various methods including needle insertion and acupressure. We include all types of acupuncture treatment, for example, manual acupuncture, electro-acupuncture, combined acupuncture with moxibustion, acupuncture combined with lifestyle management (such as exercise, control diet), or acupuncture combined with a basic treatment (such as Diane-35, Clomiphene).There will be no restrictions in terms of the needle materials, frequency of treatment sessions, and treatment courses.

##### 2.2.4 Comparison intervention

Eligible comparators were sham acupuncture, basic treatment, placebo, no treatment, lifestyle management.

#### 2.2.4 Types of outcome measures

Primary outcomes will include screening scales for anxiety or depression, such as the self-rating anxiety scale (SAS),self-rating depression scale (SDS),Hamilton rating scale for anxiety (HAMA), Hamil-ton rating scale for depression (HAMD), Montgomery Åsberg Depression Rating Scale(MADRS-S), Brief Scale for Anxiety (BSA-S).

Secondary outcomes will consider screening scales for quality of life, such as the Short-form 36 item health survey questionnaire(SF-36), Health-related quality of life questionnaire for PCOS(PCOSQ).Adverse events related to acupuncture.

### 2.3 Search strategy

The following databases will be searched from inception to July 31, 2022: PubMed, EMBASE, Web of Science, Cochrane library, China National Knowledge Infrastructure (CNKI), Wan-Fang database, Chinese Scientific Journal Database (VIP),Chinese Biomedical Literature Database (CBM). In addition, we searched Google Scholar and the ChiCTR clinical trial registration plat-Form for any relevant ongoing or unpublished trials.

Two reviewers will independently search the studies. Any differences will be resolved through discussion with a third author. The search strategy that will be run in the PubMed and adjusted to fit the other database when necessary is presented in Table.

### 2.4 Data collection and analysis

The search results will be imported into NoteExpress. After removing duplicate records, titles and abstracts will be checked to identify applicable studies by two independent reviewers. Then theyindependently extracted the data using a predesigned form.The name of the author, year of publication, inclusion and exclusion criteria, number of patients, method of randomization, allocation concealment, blinding, type of acupuncture, treatment used in the control group, outcome measures and adverse events were recorded. Any discrepancy in the process of cross-checking will be resolved by consensus. The selection process will be showed in Figure.

**Figure 1:**
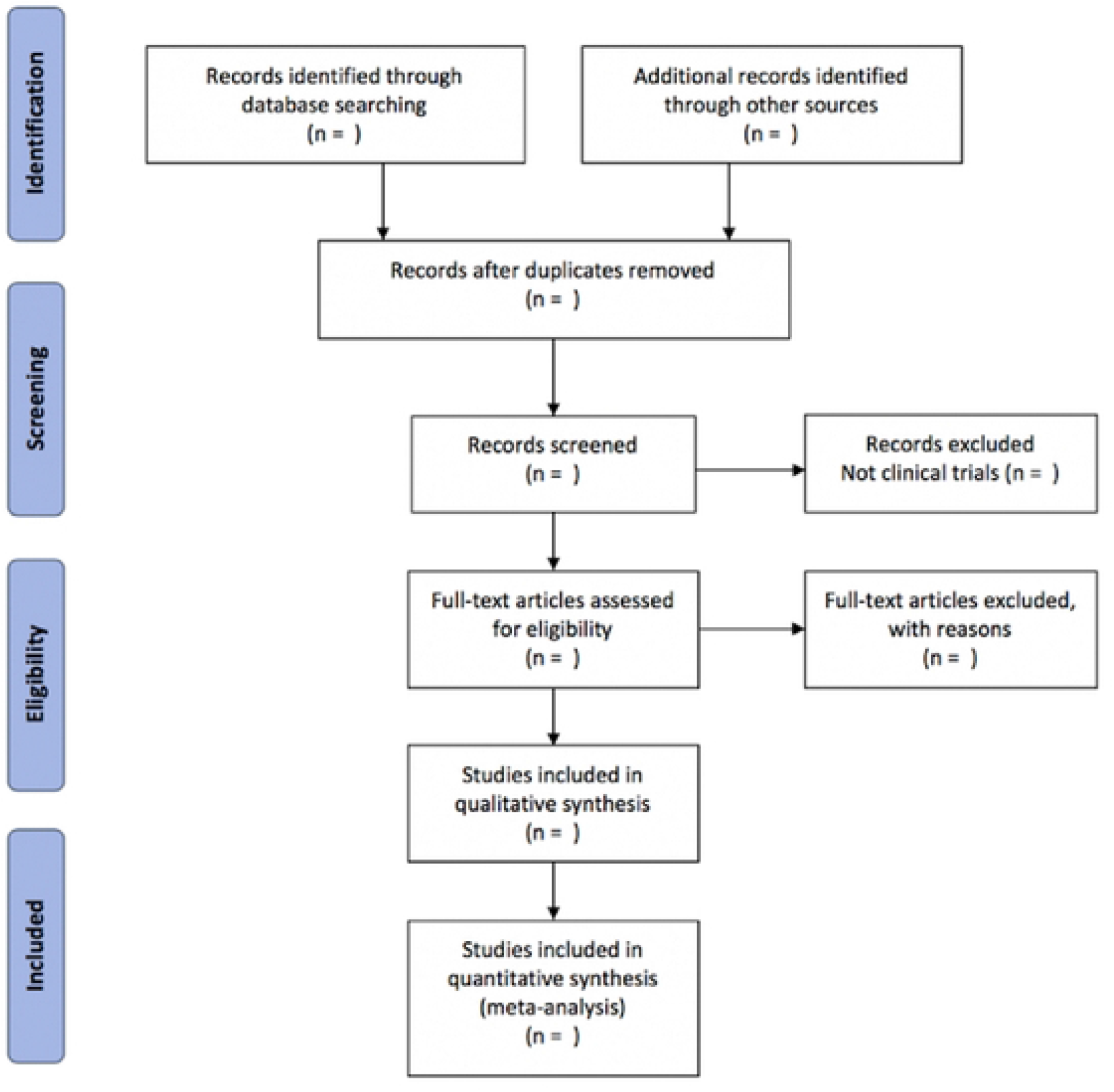
The PRISMA flow diagram of the study selection process.

### 2.5 Quality evaluation

The included studies will be evaluate according to the risk of bias assessment tools recommended by Cochrane, including random sequence generation, allocation hiding, blinded implementation, blinded outcome assessment, integrity of outcome data, selective reporting, and other bias. And we will rate the literature as “low risk”, “medium risk” and “high risk”.A third investigator will resolve any disagreements.

### 2.6 Statistical analysis

RevMan 5.4 software will be used for statistical analysis. The primary outcome measure is the score on the depression or anxiety scale after treatment. The heterogeneity of the studies is evaluated using theχ2 test and I2 statistic.Fixed-effect and random-effect models are used for the meta-analyses.We use the estimates of the random-effects model when high heterogeneity (I2 ≥ 50% or p<0.1) is present; otherwise, we use the fixed-effect model estimates. A funnel plot, as well as statistical tests (Egger test and Begg test by Stata V.15.0 software), will be used to assess reporting bias if more than 10 trials are included [25-26].

### 2.7 Subgroup analysis

If necessary, we will conduct subgroup analysis according to different types of interventions and observation indicators. If there is still significant heterogeneity, the source of heterogeneity will be explored and trials will be further classified for subgroup analysis.

### 2.8 Sensitivity analysis

If there is significant heterogeneity, we will perform sensitivity analysis for primary outcomes to check the robustness of conclusions by excluding studies with high risk of bias.

### 2.9 Grading the quality of evidence

The GRADE system will be used for evaluating the quality of evidence in systematic reviews. The evaluation included bias risk; heterogeneity; indirectness; imprecision; publication bias. The quality of evidence for RCTs will be graded as 4 levels as follow: very low, low, moderate, or high.

## 3. Discussion

This systematic review is expected to provide reference for acupuncture in the treatment of affective symptoms of PCOS.We hope the results of this study will provide guidance for physician’s decision-making and the development and update of guidelines.

Patient and public involvement statement No patient involved.

## Data Availability

No datasets were generated or analysed during the current study. All relevant data from this study will be made available upon study completion

## Ethics and dissemination

This review will not involve private information from individuals and will not affect patient rights therefore does not require ethical approval. The results of this review will be disseminated through peer-reviewed publications and conference reports.

## Data Availability

All data are fully available without restriction. All relevant data are within the paper and its Supporting Information files.

